# Non-obese lipedema patients show a distinctly altered Quantitative Sensory Testing profile with high diagnostic potential

**DOI:** 10.1101/2023.04.25.23289086

**Authors:** Rebecca Dinnendahl, Dominik Tschimmel, Vanessa Löw, Manuel Cornely, Tim Hucho

**Author notes:** Corresponding author: Tim Hucho Mail to,Translational Pain Research, Department of Anaesthesiology and Intensive Care Medicine University Hospital Cologne, University Cologne Joseph-Stelzmann Str 9, 50931 Cologne, Germany. **Disclosures:** The authors have declared that no conflict of interest exists.

## Abstract

**Background:** Lipedema is a widespread severe chronic disease affecting mostly women. Characterized by painful bilateral fat accumulation in extremities sparing hands and feet, objective measurement-based diagnosis is currently missing. We tested for characteristic psychometric and/or sensory alterations including pain and for their potential for medical routine diagnostic.

**Methods:** Pain-psychometry were assessed using the German Pain Questionnaire. Sensory sensitivity toward painful and non-painful stimuli was characterized in non-obese lipedema patients and matched controls using the validated QST-protocol of the German Research Network on Neuropathic Pain.

**Results:** Lipedema patients showed no overt psychometric abnormalities. Pain was reported as somatic rather than psychosomatic-aversive. All QST measurements were normal, but the pressure pain threshold (PPT) was twofold reduced and the vibration detection threshold (VDT) was two and a half times increased. Both thresholds were selectively altered at the affected thigh but not the unaffected hand. ROC-analysis of the combination of PPT and VDT of thigh versus hand into a PVTH-score shows high sensitivity and specificity, categorizing correctly 96.5% of the participants as lipedema patients or healthy controls. Bayesian inference analysis corroborated the diagnostic potential of a combined PVTH score.

**Conclusion:** We propose to assess PPT and VDT at the painful thigh and the pain-free hand. Combination in a PVTH-score may allow a convenient lipedema diagnosis early during disease development.

**Trial Registration:** German Clinical Trials Register (DRKS00030509)

**Funding:** The project was funded by the DFG (459479161).

## Introduction

Lipedema, also known as Lipohyperplasia dolorosa (LiDo) is a widespread bilateral subcutaneous deposition of adipose tissue in limbs and arms not affecting feet or hands (1–12). Depositions are unresponsive to dietary restrictions or physical activity (5,6,13). Lipedema affects almost exclusively women and typically manifests concomitant with hormonal changes, such as puberty, pregnancy, or childbirth (14,15).

Pain is considered a lipedema-defining characteristic (1,16–21). It is perceived in the affected extremities and differentiates lipedema from non-painful phenotypes such as obesity or lymphedema (19). The etiology of lipedema pain is currently unknown. Patients are mostly unresponsive to analgesics and this lasting pain greatly aggravates the burden of the disease (16,18).

Lipedema pain is ill described. It has been described as sensitivity against touch but also as continuous pain. It is described as “if legs would burst from the inside”, “painful weaknesss”, “piercing, stabbing” (22). It remains unclear, which of the clinical pain categories such as nociceptive, inflammatory, neuropathic, or psychosomatic pain may be at the heart of this debilitating condition. An objectifiable characterization of lipedema pain beyond patient-self-reporting is currently missing. Pain is defined as a physiological-sensory and psychological-emotional experience (23). The emotional experience of lipedema pain is routinely recorded by pain questionnaires such as “Deutsche Schmerzfragebogen” or “painDETECT” (2,24,25). In contrast, it has not been attempted to characterize, which physiological sensory sensitivities such as e.g. detection of warmth, cold, heat pain, cold pain, pressure pain may be changed and to quantify such changes in lipedema patients.

Accordingly, we now aimed to characterize the somatosensory phenotype in lipedema-patients using the standardized approach of quantitative sensory testing (QST) as developed by the German Research Network on Neuropathic Pain (DFNS) (26–29). Conducting 7 tests, 13 different sensory thresholds are determined. Objectivity was assured by averaging over repetitive tests, standardized training of the measuring personal, comparison of control measurements to over 1000 database controls, comparing lipedema patients with unaffected matched controls, as well as measuring the unaffected hands in addition to the affected thigh, which served as patient-specific internal control. Finally, yet importantly, we focused on young non-obese patients, which remain largely undiagnosed for decades. The study was accompanied by a standard pain questionnaire used in Germany to investigate patients’ psychometry and pain descriptions to provide a comprehensive analysis of the hallmarks of lipedema pain. The potential of the results for differentiating lipedema from controls was tested by ROC-analysis and confirmed by Bayesian inference analysis.

## Results

### Study population consisted of non-obese age- and WtHR-matched women with only minor comorbities

We recruited 20 women per group. The study was conducted in German with all 40 participants speaking German on native speaker level (for overview of cohort characteristics see table 1). There was no statistically significant difference of age (ctrl: 27.15 ± 4.2 years, lipedema: 27.35 ± 4.4 years; *p* = n.s), height (ctrl: 169.3 ± 6.0 cm, lipedema: 165.7 ± 7.3 cm; *p* = n.s.), weight (ctrl: 63.4 ± 7.7 kg, lipedema: 68.3 ± 11.1 kg; *p* = n.s.), waist (75.2 ± 5.1 cm, lipedema: 76.3 ± 7.8 cm; *p* = n.s.), and waist to height ratio (WtHR, waist[cm]/height[cm], ctrl: 0.44 ± 0.03, lipedema: 0.46 ± 0.04; *p* = n.s.). lipedema patients showed a slight but statistically significant higher body-mass index (BMI, weight[kg]/(height[m])²) compared to the controls (ctrl: 22.1 ± 2.4 kg/m², lipedema: 24.8 ± 2.9 kg/m², *p* < 0.05). BMIs and WtHR of both groups were within the normal or slightly over-weight range (30) (see figure 1).

**Figure 1:**
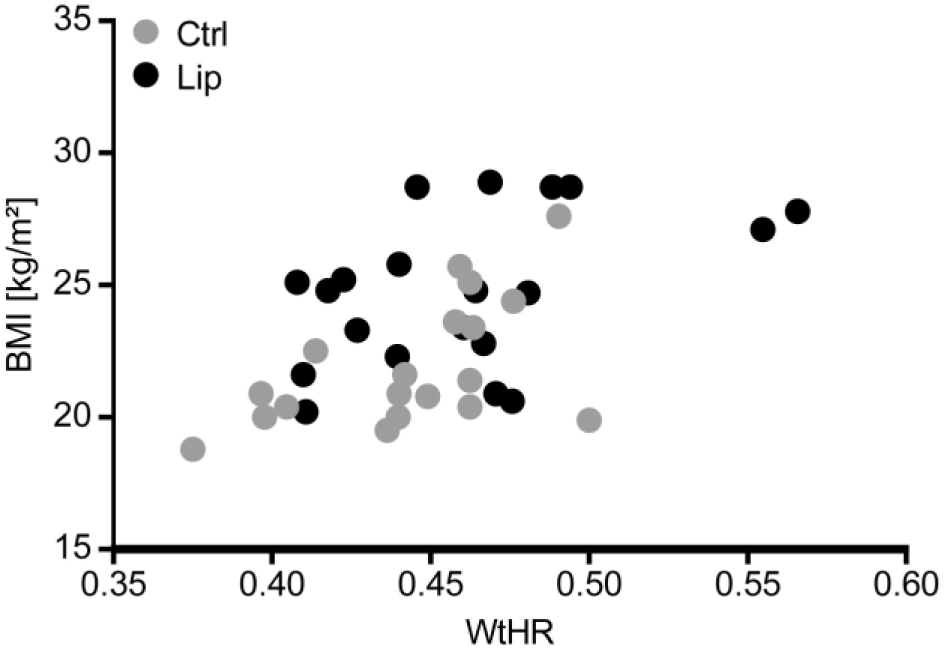
Distribution of BMI and WtHR of our study population. Lipedema (Lip) patients showed a slight but statistically significant higher body-mass index (BMI) compared to the controls. Waist-to-Height-Ratios (WtHR) of both groups were not significantly different. BMIs and WtHR of both groups were within the normal or slightly over-weight range.

**Table 1:** Biometrical data.

|  | **Ctrl** |  | **Lipedema** |  | ***p*** |
| --- | --- | --- | --- | --- | --- |
|  | Mean +/- SD | range | Mean +/- SD | range |  |
| age [years] | 27.15 +/- 4.2 | 20 - 37 | 27.35 +/- 4.4 | 23 - 40 | n.s. |
| height [cm] | 169.3 +/- 6.0 | 157 - 179 | 165.7 +/- 7.3 | 152 - 175 | n.s. |
| weight [kg] | 63.4 +/- 7.7 | 48 - 76 | 68.3 +/- 11.1 | 54 - 88 | n.s. |
| waist [cm] | 75.2 +/- 5.1 | 60 - 80 | 76.3 +/- 7.8 | 66 - 99 | n.s. |
| WtHR | 0.44 +/- 0.03 | 0.38 - 0.5 | 0.46 +/- 0.04 | 0.41 - 0.57 | n.s. |
| BMI [kg/cm<sup>2</sup>] | 22.1 +/- 2.4 | 18.8 - 27.6 | 24.8 +/- 2.9 | 20.2 - 28.9 | < 0.05 |

Psychometric parameters and comorbidities were assessed using the DSF questionnaire. While all received the DSF, only the 14 lipedema patients providing the full information were analyzed.

All lipedema patients were diagnosed as stage I or II (6) at least 6 months before the measurement (11.2 ± 6.6 years, range 0.5 – 27 years). They associated the manifestation of the disease with phases of hormonal changes, such as puberty, and 14 reported a familial history of lipedema with affected kin. All lipedema patients reported perceived chronic pain in the affected legs and in 85.7% of patients the pain was present for 1 year or longer. All participants reported only minor comorbidities (see table 2).

**Table 2:** Comorbidities.

| **Comorbidities** | **Ctrl (n = )** | **Lipedema (n = )** |
| --- | --- | --- |
| Mental/ emotional strain | 2 | 2 |
| Hypothyreosis | 3 | 2 |
| Asthma | 1 | 0 |
| Migraine | 0 | 1 |
| Chronic sinusitis | 0 | 1 |
| Reflux, gastritis | 2 | 1 |
| Focal nodular hyperplasia | 1 | 0 |
| Endometriosis | 0 | 2 |
| Orthopedical entities (scoliosis, backpain, ligament rupture) | 1 | 3 |
| Peripheral nerve injury (area out of interest) | 1 | 0 |

### Lipedema patients showed no signs of depression, anxiety, or stress and lacked indications for concerning mental abnormalities

The DSF-questionnaire includes the “Depression, Anxiety, Stress Scale (DASS)”. All scores for both groups were in an asymptomatic range i.e. below threshold of clinical significance (dashed red lines) (Figure 2A). Nevertheless, all scores were significantly higher in lipedema patients compared to controls with respect to depression (controls 2.4 ± 3.66 versus lipedema 5.57 ± 4.26, *t*(32) = 2.33, *p* < 0.05), anxiety (controls 1 ± 1,3 versus lipedema 2.86 ± 3.03, *t*(32) = 2.45, *p* < 0.05), and stress (controls 3.1 ± 2.47 versus lipedema 7.29 ± 4.34, *t*(32) = 3.58, *p* < 0.01).

**Figure 2:**
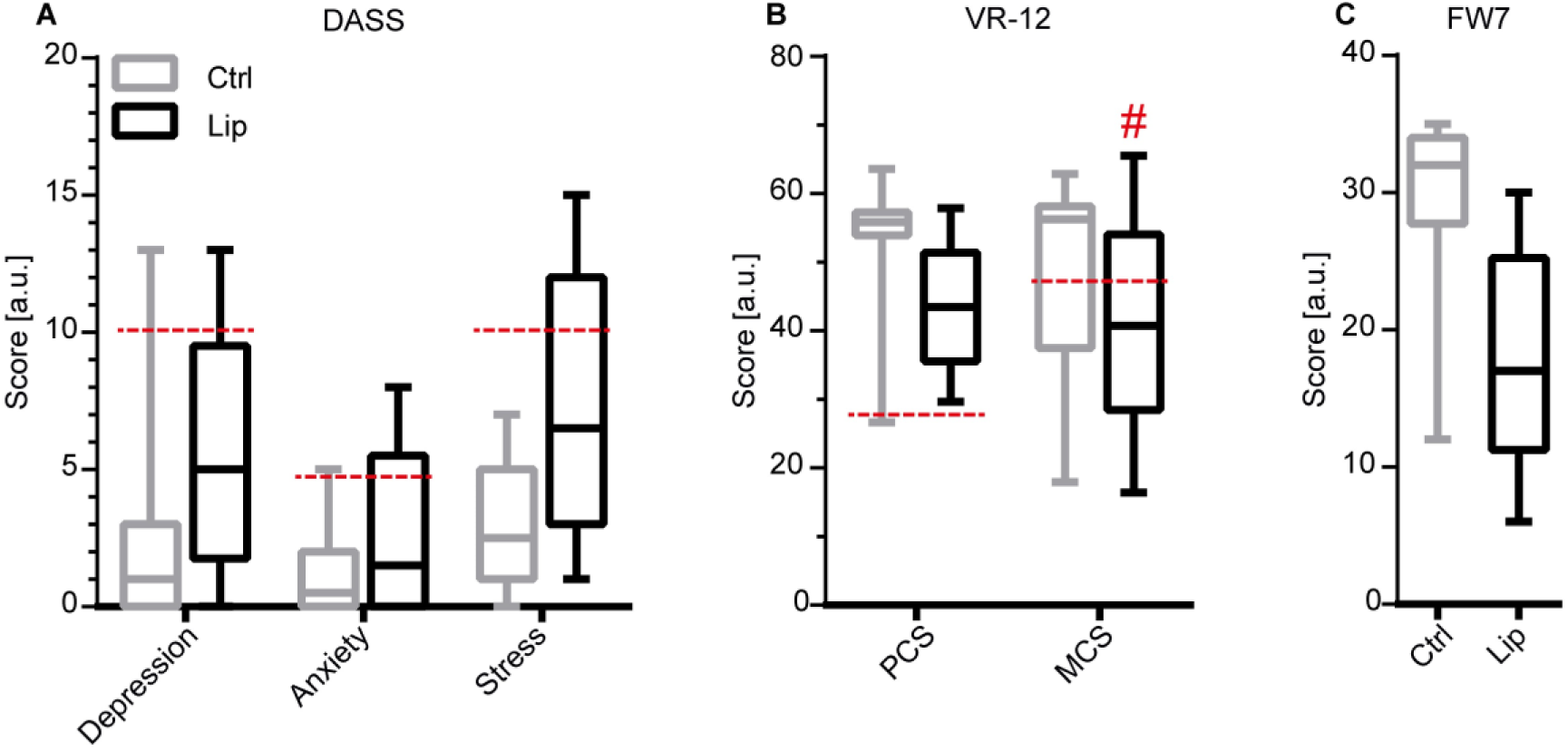
Psychometry of the participants as measured by the DSF. Dashed lines indicate cut-off values separating scores considered as normal or abnormal, respectively. (**A)** All scores of the depression-anxiety-stress scale (DASS) remained below the cut-off values and thus are considered as normal. **(B)** Results for the general health condition (veterans RAND-12 (VR12)) questionnaire with respect to the “physical compartment summary (PCS)” and “mental compartment summary (MCS)”. Scores above dashed lines are considered as normal values. We found normal scores for both groups in the PCS, MCS scores slightly below the threshold value in lipedema (Lip) patients indicating the presence of minor mental burden. **(C)** Results for the habitual well-being (FW7 questionnaire) with higher scores indicating more well-being. We found a reduced score in lipedema patients; however, still in the mid-range of the scale, indicating normal habitual well-being values for patients with chronic pain. (All values are displayed as mean + standard deviation. Ctrl n = 20, lipedema n = 14).

General health condition was assessed using the VR12 as part of the DSF-questionnaire. The score is subdivided into a “physical compartment summary (PCS)” and a “mental compartment summary (MCS)”. PCS-Scores of both groups were asymptomatic (values above dashed red line, Figure 2B). While non-pathological, the lipedema group showed reduced scores (43.78 ± 8.69) compared to controls (54.25 ± 7.69), *t*(30) = 3.59, *p* < 0.01. For MCS, the lipedema group showed slightly symptomatic values being below the cut-off value of 43. Nevertheless, we did not find a significant difference between the groups (Ctrl: 48.38 ± 14.56, lipedema: 40.99 ± 15.46), *t*(30) = 1.38, *p* > 0.05).

Furthermore, the DSF-questionnaire assess the habitual well-being using FW7 (see figure 2C). Higher scores indicating higher well-being. Without a clear cut-off value, scores from the mid-range and up can be considered as normal. The lipedema group scored mid-range (17.64 ± 7.84) and controls higher range (29.94 ± 5.81), *t*(30) = 5.1, *p* < 0.0001) Both scores indicate normal habitual well-being in both cohorts.

### Lipedema patients report severe persistent pain with circadian fluctuations described with somatic terms

All participants rated their pain intensity on a numerical rating scale (NRS) during resting as well as during stress such as mild exercise (figure 3A). Control participants did not report noticeable pain with the exception of two participants with very mild stress-induced pain perceptions due to occasional non-chronic posture-induced back pain. In contrast, lipedema patients reported pronounced pain at resting conditions (6.00 ± 2.18), *t*(32) = 12.37, *p* < 0.0001, and increased stress-induced pain intensities (7.43 ± 1.91), *t*(32) = 11.96, *p* < 0.0001.

**Figure 3:**
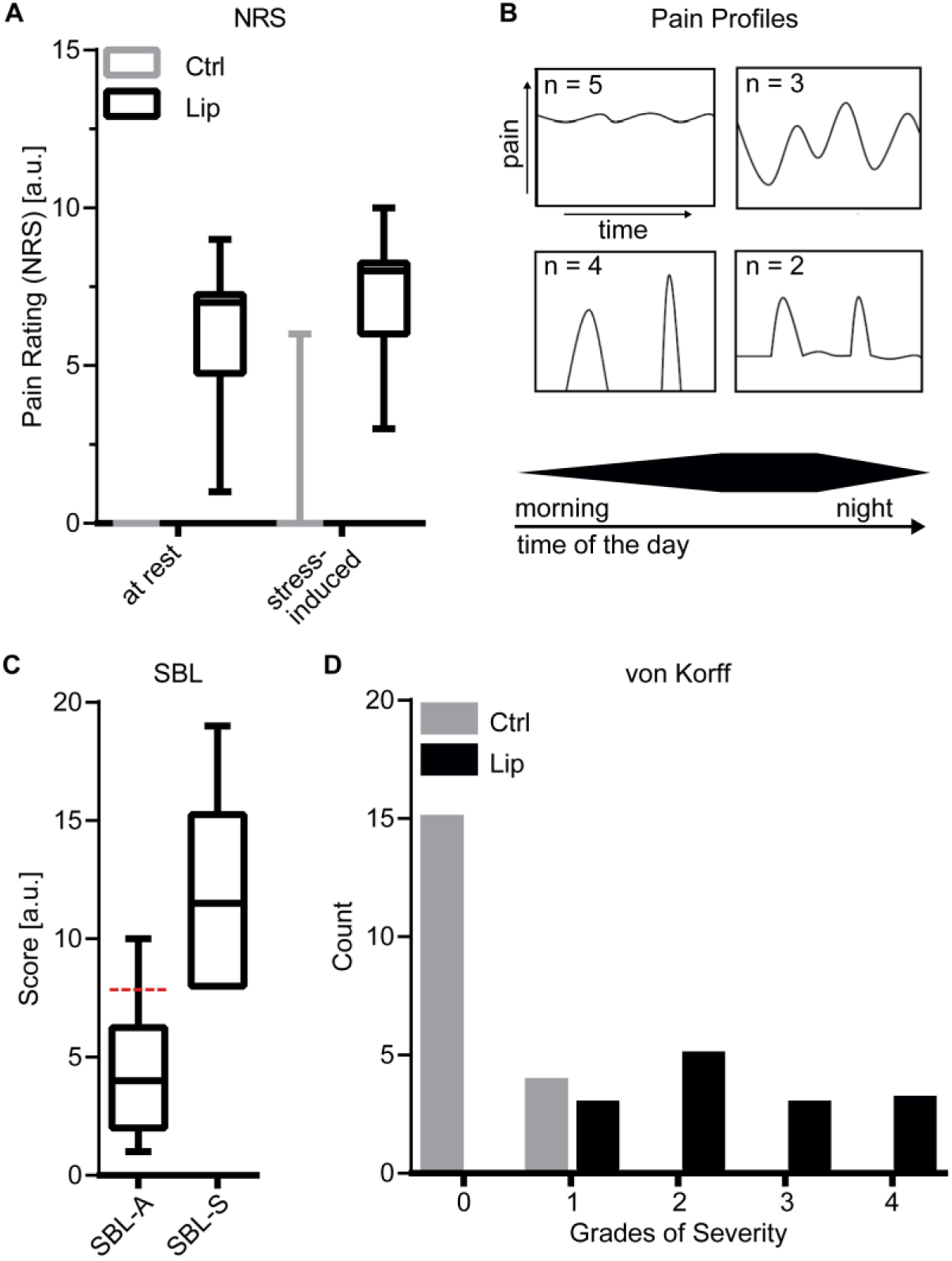
Characterization of Lipedema (Lip) pain as measured by the DSF. **(A)** Pain intensity ratings on numerical rating scale (NRS;0 = no pain, 10 = worst imaginable pain) under resting conditions and stress-induced, e.g. during mild exercise. Lipedema pain ratings were significantly increased compared to the control group, where pain was virtually absent. (Ctrl n = 20; lipedema n = 20; independent t-test; **** *p* < 0.0001). **(B)** Pain profiles (modified from [10]) as described by lipedema patients with circadian fluctuations. **(C)** Results for the German version of the Pain Description List (SBL), subdivided into an affective (SBL-A) and somatic (SBL-S) part. Values above the dashed line indicate a pathologic SBL-A of increased affective pain perception. This was not the case in our population of lipedema patients. (n = 14). Furthermore, the higher SBL-S score indicated a rather somatic nature of lipedema pain. **(D)** Grades of severity according to von Korff (0: no pain; 1: low pain intensity; low disability; 2: high pain intensity; low disability; 3: high pain-related disability; moderately limiting; 4: high pain-related disability; severely limiting). (Ctrl n = 20; lipedema n = 14; chi square test; **** *p* < 0.0001).

All lipedema patients reported a distinct circadian pattern with increasing pain, starting at around early afternoon and culminating in the evening (figure 3B). The pain was experienced very individually with various degrees of oscillation and/or attacks. All but 4 reported continuous pain.

We used the pain description list (Schmerzbeschreibungsliste (SBL) (31) to capture the emotional or affective part (SBL-A) and the somatic part (SBL-S), respectively (see figure 3C). SBL-A values remained considerably below the threshold value. SBL-S presented higher values. This indicated a subordinated role for the affective emotional component, while pointing to a rather somatic nature of lipedema pain.

The von Korff grading captures the severity of pain as a function of intensity and disability (32) (see figure 3D). Grades are defined as 0: no pain, 1: low pain intensity and low disability, 2: high pain intensity with low disability, 3: high pain-related disability that is moderately limiting, and grade 4: high pain-related disability that is severely limiting. Corroborating others, lipedema pain appears in average as moderately in extreme cases though also as severely limiting.

### Normal sensitivity thresholds for all lipedema patients and controls measured at the dorsum of the hand

Going beyond questionnaire-based psychometry, we performed QST according to the protocol of the DFNS (26–29) to objectify evoked response thresholds of sensory inputs.

First, sensory thresholds were assessed at the non-affected dorsum of the hand. Comparison with DFNS control data confirmed threshold z-scores for all parameters to remain in the normal range within the 95% confidence interval (CI) (-1.96 to 1.96). A repeated measures ANOVA followed by Sidak’s post hoc test showed no significant difference between both groups in any of the parameters assessed at the dorsum of the hand (figure 3a), F(1, 418) = 0.0002, *p* > 0.05. This indicates experimenter-proficient using the QST-methodology and thus absence of generalized pain (figure 3A)

### Selectively decreased threshold for pressure pain and increased threshold for vibration detection at the lateral thigh of lipedema patients

Next, measurements were conducted at the lateral thigh as the area with reported pain sensation in patients (see figure 4B). Z-scores of the control group remained within the normal 95% CI range, with exception of a slightly increased value for the pressure pain threshold if compared to DFNS control measurements performed at the dorsum of the foot. Also, lipedema patients showed normal QST measurements for most test stimuli with two exceptions: 1) values for the PPT were strongly increased (4.51 ± 1.26, see figure 4C), indicating pain hyper-responsiveness; 2) values for the VDT were strongly decreased (-3.67 ± -1.41, figure 4C) suggesting reduced sensitivity to vibration. Repeated measures ANOVA followed by Sidak’s multiple comparison post hoc test revealed a significant difference between lipedema patients and controls (F(37, 370) = 2.485, *p* < 0.0001) in the PPT (p < 0.0001, 95% CI = -3.442, -1.371) and the VDT ((p < 0.0001, 95% CI = 1.203, 3.274).

**Figure 4:**
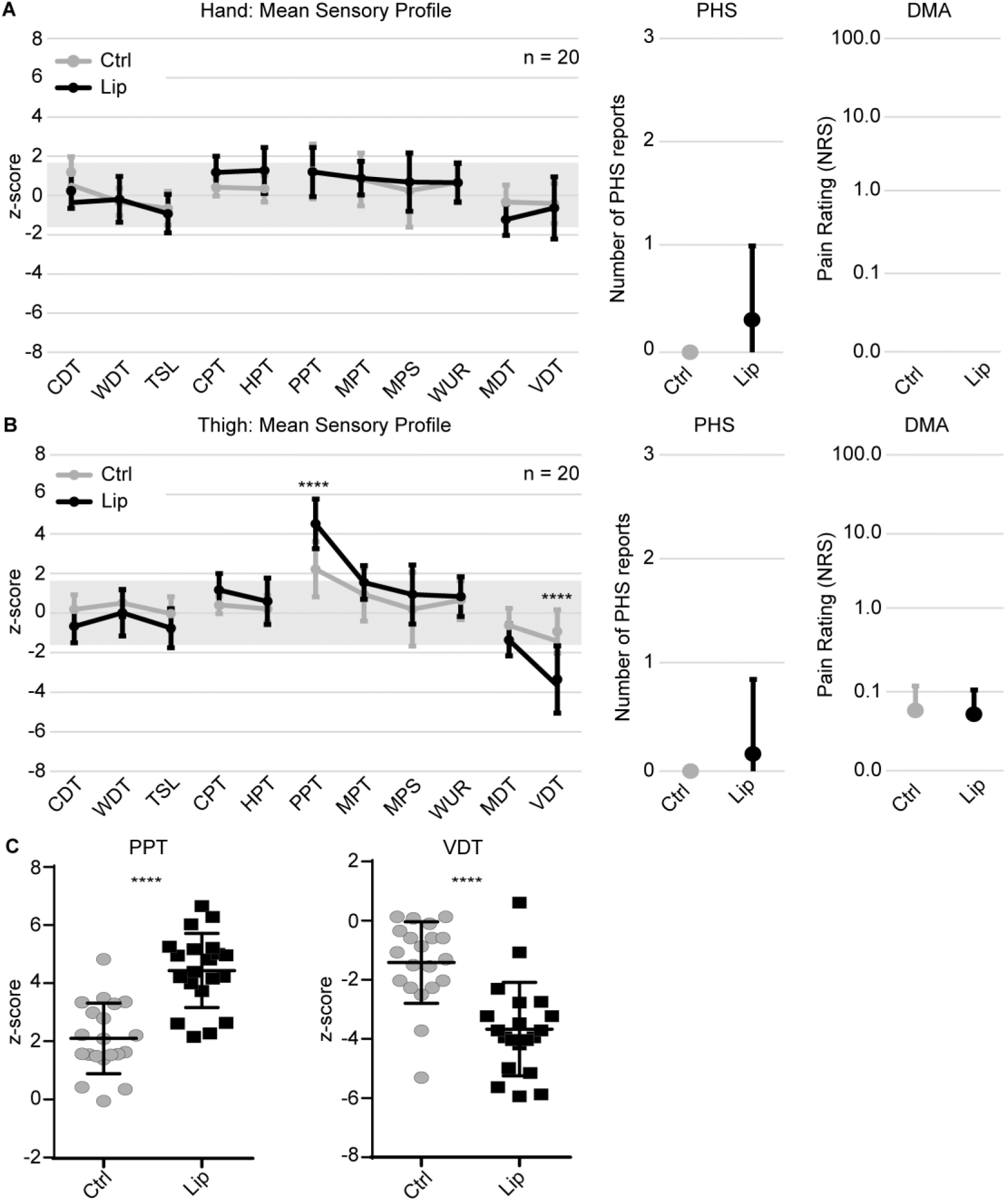
Mean QST Sensory Profiles. **(A)** Mean QST sensory profiles of control and lipedema (Lip) participants measured at the dorsum of the hand. Values between -1.96 and 1.96 are considered normal. **(B)** Mean QST sensory profiles of control and lipedema participants measured at the lateral thigh. We found significantly increased PPT and decreased VDT values, respectively, in lipedema patients **(C)** Display of single participant data of controls and lipedema patients measured at the lateral thigh for PPT and VDT. (CDT cold detection threshold, WDT warmth detection threshold, TSL thermal sensory limen, CPT cold pain threshold, HPT heat pain threshold, PPT pressure pain threshold, MPT mechanical pain threshold, MPS mechanical pain sensitivity, WUR wind-up phenomenon, MDT mechanical detection threshold, VDT vibration detection threshold, PHS paradoxical heat sensations, DMA dynamical mechanical allodynia). (Ctrl n = 20, lipedema n = 20 (except thermal thresholds at the lateral thigh: n = 19 (see results section for explanation)), two-way repeated measures ANOVA, **** *p* < 0.0001).

### Assessment of PPT or VDT shows high sensitivity and selectivity to identify participants as lipedema patients

Next, we investigated whether consideration of only the two altered parameters allows for a reliable reassignment of all 40 measured women as either lipedema patient or normal-control. For this, a ROC analysis for sensitivity and specificity of such an assignment was performed. First, we tested if using either the values for the PPT or alternatively for the VDT would allow to correctly identify participants as either lipedema patient or control. Each parameter alone showed promising diagnostic ability to distinguish lipedema and control participants, assigning in the best case 90.75 % (PPT) and 86.38 % (VDT) of the measured women correctly as lipedema or control (PPT: AUC = 0.9075, *p* < 0.0001; VDT: AUC = 0.8638, *p* < 0.0001, figure 5A).

**Figure 5:**
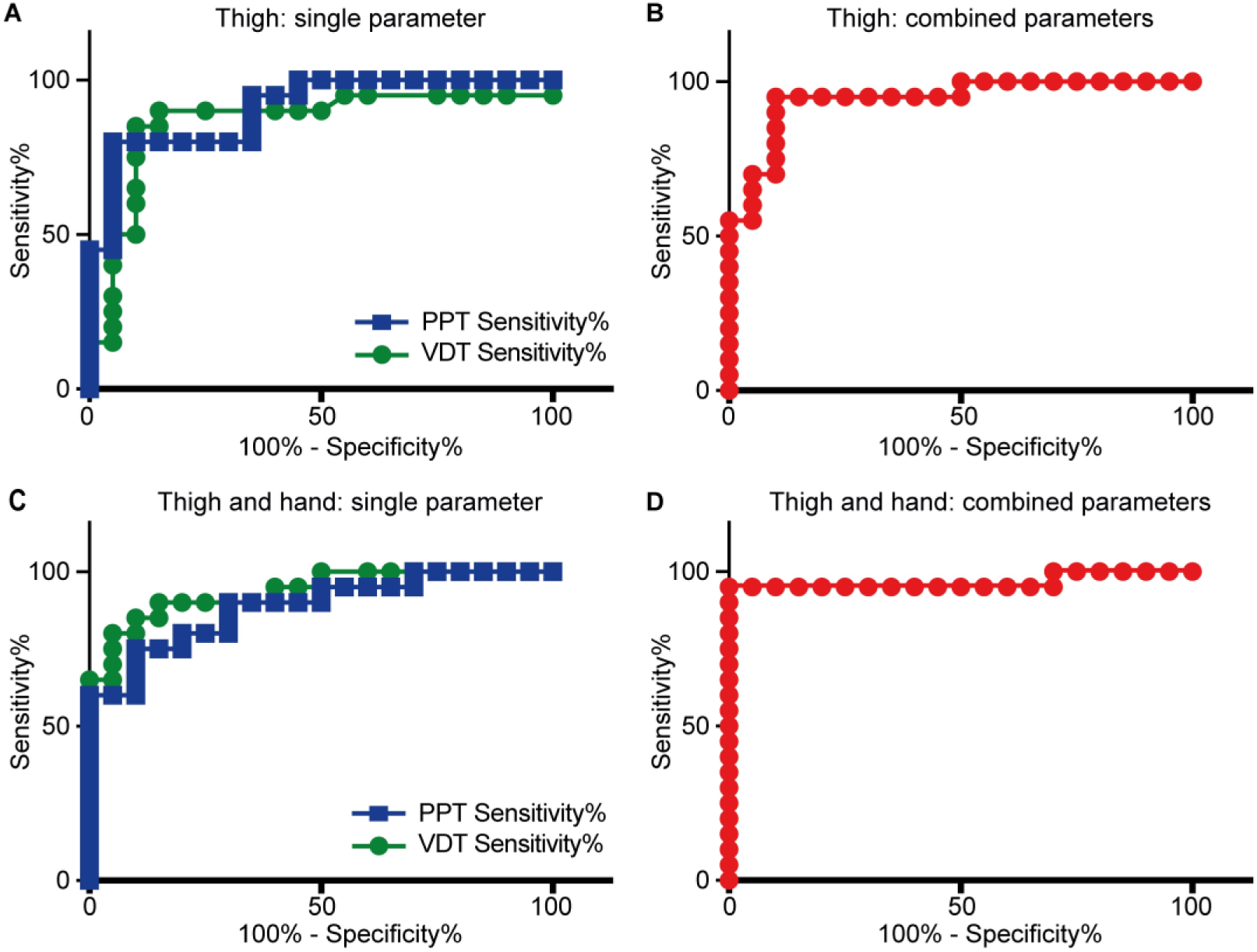
ROC analyses for diagnostic ability investigation of assessed QST z-scores. **(A)** ROC analyses of PPT and VDT measured at the lateral thigh in control participants and lipedema (Lip) patients. Each parameter alone showed promising diagnostic ability to distinguish both groups of our study population. (**B)** ROC analysis of the sum absolute values of both parameters on single patient level. Assessment of both parameters increased the diagnostic ability. **(C)** Intraindividual control measurements are considered by absolute value subtraction of hand measurements from measurements of the thigh for each parameter. Again, both parameters showed promising diagnostic ability. **(D)** addition of both values calculated in c) showed the highest diagnostic potential in terms of sensitivity and specificity.

### Combination of PPT and VDT values shows higher sensitivity and selectivity to assign participants as lipedema patients

Next, we asked whether combining PPT and VDT potentially allows an even better identification of single individuals as either lipedema patient or control. We summed the absolute values of the z-scores of PPT and VDT measured at the lateral thigh and performed another ROC analysis. Combining both parameters increased the diagnostic ability to even 94.25 % correct assignment as lipedema or control (AUC: 0.9425, *p* < 0.0001, figure 5B).

Since we measured all QST parameters at the hand as an intra-individual control site, we tested whether a combination of the QST measurements taken at the thigh with the ones taken at the hand allows an even more sensitive and selective group assignment. For this, we subtracted the absolute z-score hand-values from the respective thigh-values of the same individual for PPT as well as separately for VDT, respectively (Δ_(parameter)_ = z-score_(thigh)_ – z-score_(hand)_, *d*-score). This did not increase the sensitivity and selectivity to assign measured women as lipedema or control (figure 5C, PPT: AUC 0.885, *p* < 0.0001; VDT: 0.935, *p* < 0.0001).

### Integration of all 4 measurements (PPT-thigh, PPT-hand, VDT-thigh and VDT-hand,) into a PVTH-score shows best sensitivity and selectivity to identify participants as lipedema patients or controls

Last, we combined all four measurements into a PVTH-score and tested for its sensitivity and selectivity to identify the measured women as lipedema or control. The combined score was defined by us as PVTH-score=Δ_(PPT)_ - Δ _(VDT)_. Of all ROC analyses this resulted in best sensitivity and best specificity, identifying 96.5 % of the measured individuals correctly as lipedema patient or as control (AUC = 0.965, *p* < 0.0001, figure 5D, table 3 provides a list with respective sensitivity-specificity pairs for exemplary criterion values).

**Table 3:**
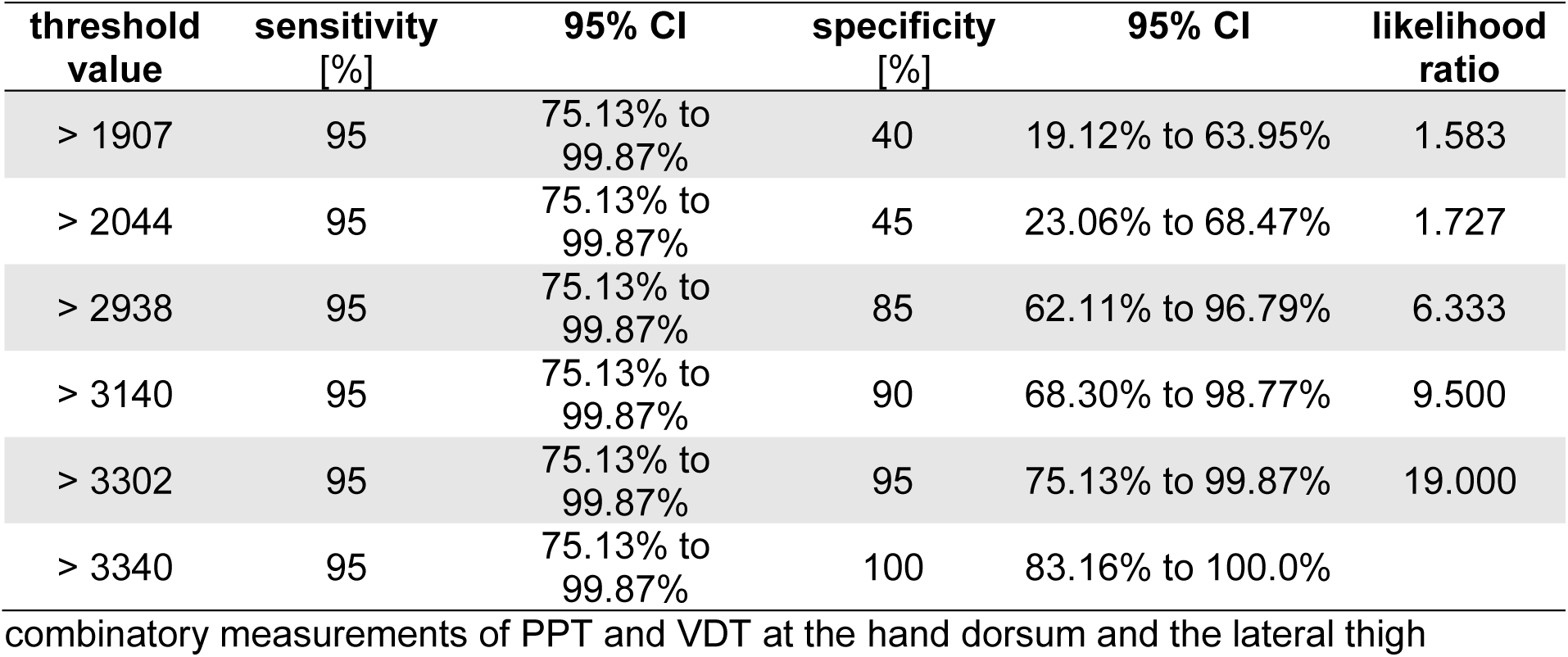
Exemplary sensitivity-specificity threshold value pairs.

Taken together, we conclude that a joint assessment of z-scores of only PPT and VDT at the dorsum of the hand and the lateral thigh (PVTH-Score) shows promising power for the differentiation of lipedema versus control. This suggests that one may reduce the full QST-protocol of 7 measurements at two different sites to just these two measurements thereby reducing the time from about 1-1.5 h for a full QST to about 10 minutes for a just two measurement protocol while maintaining a high sensitivity and selectivity for the identification of lipedema patients on a single patient basis.

### Bayesian inference confirms promising diagnostic ability of 4 combined measurements (PPT-hand, PPT-thigh, VDT-hand, and VDT-thigh) irrespective the cohort size of the study on hand

Transferability of studies depends on the investigated cohort. Especially for small sample sizes classical statistics does not provide reliable estimates for generalization of results. In contrast, Bayesian statistics can quantify parameters for generalization to larger populations by providing the probability that a particular parameter is the true but unknown parameter of the general population. With this, we can estimate how well our proposed diagnostic test would perform in the medical practice.

For the Bayesian analysis we considered the 𝑑-scores from 19 lipedema patients and 20 non-lipedema participants (figure 6A). How the 𝑑-scores for the whole population are distributed is not known. For the general population estimate we assume a location-scale 𝑡-distribution 𝑡(𝜇, 𝜎, 𝜈) for the 𝑑-scores of the general population. The parameters 𝜇, 𝜎 and 𝜈 have to be inferred from our small data set {𝑑_𝑖_}. For this, the following posterior probability distribution is used:

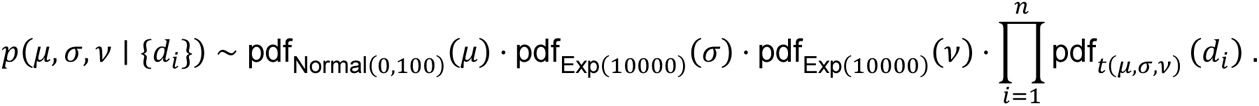

**Figure 6:**
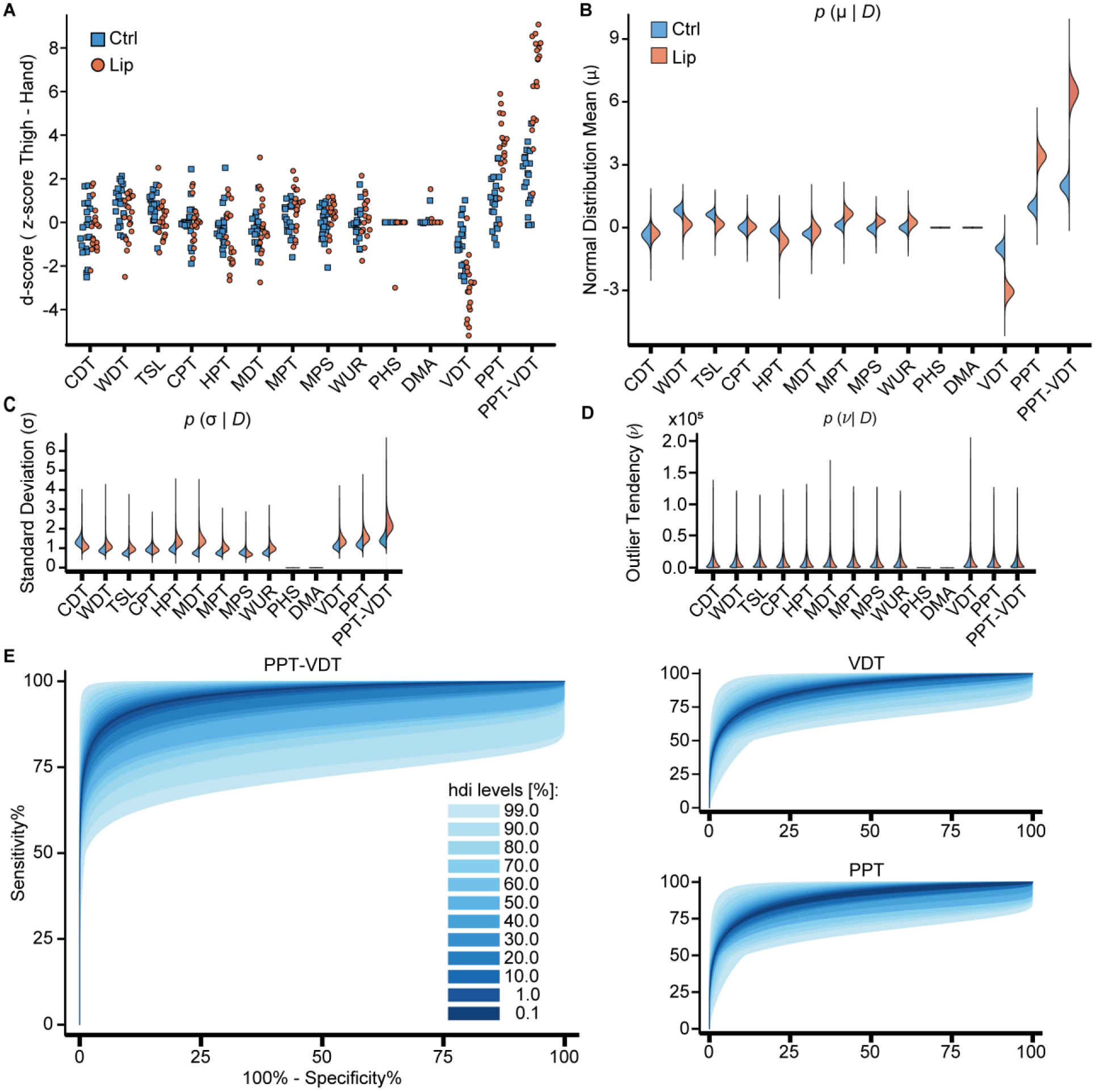
Bayesian inference about the general population from small sample size. **(A)** difference of z-scores from lateral thigh and hand, named “d-scores”, for the different measurements of control and lipedema (Lip) women. **(B), (C), and (D)** Inferred parameters for both groups for all QST-aspects. The combined PPT-VDT 𝑑-scores difference between lipedema and non-lipedema is more pronounced than PPT or VDT alone (𝜇 :mean of a normal distribution, 𝜎 :standard deviation and 𝜈 :outlier tendency). **(E)** ROC curve for PPT-VDT calculated from the inferred population distributions. Color shades display range of possible ROC curves for different highest density interval (hdi) levels. The darker the shade, the lower the corresponding hdi level. The combined 𝑑-score PPT-VDT promises to be a valid diagnostic tool with reasonable sensitivity and specificity for the detection of lipedema.

A detailed explanation and derivation can be found in the supplement. In all brevity, the posterior distribution allows calculation of the probability that a parameter is the true but unknown population parameter, on the basis of the limited data collected so far. Figure 6B-D shows these probability distributions for the individual parameter: location 𝜇, scale 𝜎 and outlier tendency 𝜈. Table 4 lists the 99% highest density intervals, i.e. the smallest intervals in which the true parameters lie with 99% probability, given our data. In addition, the posterior probability allows to plot different credibility regions for ROC curves that are to be expected if our study was repeated with other and potentially more participants (figure 6E).

**Table 4:** 99% highest density intervals for the QST parameters of interest.

| **parameter** | **VDT** | **PPT** | **PPT-VDT** |
| --- | --- | --- | --- |
| $\mu$ non-lipedema | [-1.71, -0.31] | [0.21, 1.75] | [1.09, 2.90] |
| $\mu$ lipedema | [-3.96, -2.16] | [2.40, 4.45] | [5.02, 7.91] |
| $\sigma$ non-lipedema | [0.72, 1.76] | [0.80, 1.95] | [0.93, 2.28] |
| $\sigma$ lipedema | [0.88, 2.22] | [1.02, 2.56] | [1.43, 3.60] |
| $\nu$ non-lipedema | [1.21, 46213.70] | [1.21, 46089.20] | [1.83, 45991.70] |
| $\nu$ Lip | [0.96, 46113.61] | [0.97, 45980.46] | [0.97, 46159.00] |

These analyses corroborate our results: The PVTH-score appears as a promising diagnostic test also for the general population. The credibility regions of the ROC analysis suggest that our sample ROC analysis (figure 5) can be in principle generalized to the general population. The 99% credibility levels contain as best case a sensitivity above 95% and as worst case a sensitivity of at least 50%, both for negligible false positive probabilities (100% - specificity). The most probable sensitivity (0.1% hdi region) is around 75%.

## Discussion

Pain is a hallmark reported by most lipedema patients. We aimed to objectify lipedema pain for its psychosocial versus physiological sensory experience. This may guide hypothesis building about the etiology and treatment of lipedema pain and may help developing novel diagnostic tools. Our sample size estimation resulted in a surprise. Anecdotal QST-data of clinical routine patients indicated a pronounced effect size detectable with a cohort of 17 patients. Indeed, our cohort of 20 non-obese lipedema patients and 20 matched controls corroborated the existence of a clear twofold increased PPT-z-score for lipedema patients over our matched controls. The difference to the over 1000 QST-database controls was even larger. We believe, our patients were nicely matched by weight, height, waist, and WtHR, respectively (7–10,30,33,34). Though small, they represented the general population with comorbidities such as orthopedical problems, hypothyreosis (35,36), occasional back-pain and migraine. Participants were only excluded if diagnosed as chronic pain patients but not with only anecdotal pain. We even kept two participants with endometriosis since their QST profiles did not systematically differ.

An upcoming topic is the psychological burden of lipedema patients (37,38). We do not have indications of clinically relevant psychometric abnormalities. All participants reported normal scores for depression, anxiety, stress (DASS questionnaire), showed no significant influence of stress on pain experiences, normal PCS score (VR-12 questionnaire), and a normal general well-being, except for a marginally reduced VR-12 MCS score for lipedema patients. In contrast to reports by others (20), self-reported pain of stage I and II patients was severe. Corroborating others (16,18), verbal pain description pointed to a somatic rather than psychosomatic aversive experienced pain. While psychologically asymptomatic, nevertheless, lipedema patients were considerably more burdened with lower quality of life with respect to social, mental, and physical functioning (39). This may reflect the experienced chronic pain, stigmatization, reduced self-appraisal, or self-acceptance in a beau-ideal driven society (40) often aggravated by misdiagnosis or misleading treatment advice such as obligatory weight reduction.

So far, sensory thresholds such as thermal, mechanical, pressure, or vibration have not been characterized for lipedema patients. The DFNS-QST approach assessing 13 sensory thresholds (26–29) is highly reliable as it requires standardized methodological training, controls technical quality by comparison with thousands of DFNS-QST-controls, and for lipedema patients by intra-patient comparison of affected thigh and non-affected hand. With normal hand measurements we found a slightly increased PPT-z-score at the thigh, potentially reflecting, that the DFNS-standard control is the dorsum of the foot not the thigh (26–29). Still, in lipedema patients the PPT value showed a more than twofold increase. Increased adipose tissue should rather dampen pressure transmission (41). Thus, the increased PPT may reflect an objectifiable sensitization specific to lipedema patients.

Only testing with von Frey filaments, Chakraborty and colleagues reported a dynamic mechanical allodynia (20). We did not find signs thereof even though exerting more force with von Frey filaments than Chakraborty and even though extending the testing to also brush, cotton wool, and q-tips. While Chakaraborty’s measurements contrasted with their patients’ pain experience, ours were in full accord with them.

Our results may help narrowing down mechanistic hypotheses. With pressure being mediated by small or medium diameter C- or Aδ-fibers (42) and vibration by large diameter Aβ-fibers (42) no fiber type can be specifically attributed to the observed changed QST thresholds (43–45). But, with QST measures beyond PPT and VDT appearing normal, sensory innervation, stimulus detection and transmission, as well as central integration may be normal as well. In addition, local inflammation appears unlikely in absence of mechanical or thermal hyperalgesia, and systemic drivers acting on nociceptive neurons directly may be excluded as well as otherwise thresholds should be changed e.g. in hands as well.

The large and specific changes of PPT and VDT makes it attractive to explore the diagnostic potential of such focused measurements. Indeed, our post hoc ROC analyses indicates high specificity and selectivity for detecting lipedema patients. Bayesian inference analysis supported this indicating that even under worst assumed sampling conditions, nevertheless, the PVTH-score appeared as of good diagnostic potential. Reducing the full QST-protocol to a PVTH (<u>P</u>PT, <u>V</u>DT, <u>T</u>high, <u>H</u>and)-score reduces the assessment time from over 1 hour to approximately 10 minutes. Requiring only a simple tuning fork and a pressure algometer, a PVTH-score may provide a simple, time economical, and cheap bedside test. As a practical note of caution: validity of QST measurements depends on the order of measurements (46). Therefore, first VDT and then PPT should be measured first at the hand dorsum and subsequently at the lateral thigh. Which PVTH-score value then allows best lipedema-identification is currently tested on an independent larger cohort.

Our study is limited on normal to slightly overweight lipedema patients. Whether PVTH-scores are different also in obese lipedema patients is currently under investigation. Nonetheless, PVTH-score-measurements may be of great help as normal weight patients represent the majority of women at the beginning of disease manifestation. An early diagnosis is crucial to reduce the current suffering until diagnosis.

Taken together, we found no evidence psychosomatic etiology of lipedema pain. Our data provide evidence for objectifiable somatic correlates underlying the perceived pain. Furthermore, the distinct alteration of pressure pain and vibration detection thresholds at the affected thigh but not the pain-free hand allows to propose a PVTH-score with a promising potential for lipedema diagnosis. As such a score would for the first time allow to involve pain characteristics in an objectifiable manner in the medical routine for diagnosis of lipedema, we already started to validate the score in an independent cohort.

## Materials and Methods

### Patients

This project was conducted in accordance with the declaration of Helsinki and the ICH E6 Good Clinical Practice (GCP) guidelines, approved by the ethical committees (University of Cologne (20–1594), Ärztekammer Nordrhein (2021239)), and registered at the German Clinical Trials Register (DRKS00030509). All participants provided signed informed consent prior to their inclusion.

QST measurements of lipedema patients within their clinical routine indicated a difference of z-scores larger than 1 between lipedema patients and controls. Using G*Power Version 3.1.9.6 for windows we estimated a sample size of 17 plus 3 potential drop outs (effect size 𝑑 = 1, 𝜎 = 1, 𝛼 = 0.05, and a power of 80%).

Patients were recruited via the CG Lympha clinic for surgical lymphology in Cologne (inclusion criteria: female, 18 - 40 years, body mass index (BMI) below 30 kg/m²; exclusion criteria: diseases affecting the sensory system, use of topical analgesics, diagnosis of independent pain etiologies). Lipedema was diagnosed by a trained physician. Healthy controls were addressed via flyer and email within the University Hospital Cologne and the University of Cologne.

### Quantitative Sensory Testing

QST was performed according to the protocol of the German Research Network on Neuropathic Pain (DFNS) (26–29) by DFNS-trained scientists. Seven different tests were conducted to assess 13 different parameters in a standardized manner using the official DFNS test instructions and recommended testing devices (Thermal Sensory Analyser II (TSA-II; 9 cm² thermode contact area), AlgoMed digital algometer, Medoc Main Station Version 6.4.0.22 (Medoc Ltd., Israel); standardized von Frey hairs (Optihair2-Set, MRC Systems GmbH, Heidelberg, Germany); Pin-Prick stimulators (MRC Systems GmbH, Heidelberg, Germany); Rydel-Seiffert 64 Hz tuning fork (AESCULAP OF 33, AESCULAP Surgical Instruments, B. Braun, Melsungen, Germany)). Individuals were measured at the lateral thigh as one of the areas with the greatest sensation of pain in lipedema patients and the dorsum of the hand as an intraindividual unaffected control area. Vibration detection thresholds (VDT) were assessed at the patella and the processus styloideus ulnae, respectively, and pressure pain thresholds (PPT) were measured at the quadriceps femoris muscle and the thenar eminence, respectively.

Data was processed using Microsoft Excel 2010 for windows to calculate each threshold and age-, gender-, and area-normalized z-scores using the respective DFNS-reference values. Gain of functions (GOF) were defined as z-scores above the 95% confidence interval (CI) and loss of functions (LOF) as z-scores below the 95% CI.

### Assessment of Pain intensities, Psychometry, and Medical History

All participants were asked to rate their perceived pain intensities on a numerical rating scale (NRS; 0 = no pain, 10 = worst pain imaginable) under resting conditions and stress-induced, such as perceived during mild exercise. Pain psychometry was determined by the German Pain Questionnaire (DSF) of the German Pain Association (2,24), which combines several validated scores such as “The German depression-anxiety-and-stress scale (DASS)” (47), the habitual well-being (FW7) as well as general health (Veterans RAND 12; VR-12) scores (48). In addition, it contains a comprehensive section of pain descriptions, such as occurrence, courses, duration, pain description list (Schmerzbeschreibungsliste (SBL)) (31), and grades of severity according to von Korff (32), amongst others.

### Statistics

Statistics were tested using GraphPad Prism 6 for windows. Statistical significance was assumed at a level of 𝛼 = 0.05. Biometrical and psychometric data with continuous variables were compared using independent t-tests. Categorical data were tested via contingency tables by chi-square. Z-scores of QST measurements were tested with two-way repeated measures design analysis of variances (ANOVA), followed by Sidak’s post hoc tests to correct for multiple comparisons. QST data of one patient were excluded from the statistical analysis due to thermode failure but kept in the graphical representations, since thermal thresholds did not seem to be affected in lipedema patients.

Receiver operating characteristic (ROC) curves (49) were calculated for PPT, VDT and PPT-VDT measurements to gauge their potential diagnostic value.

To estimate the certainty of our results under the premise of the sample size, we performed Bayesian inference analysis (50) using the Turing package (v0.24.1) and the AdvancedMH package (v0.7.4) for Julia 1.8.5. To evaluate the posterior densities, a Hamiltonian Monte Carlo (HMC) algorithm with No-U-Turn Sampler (NUTS) were used to obtain 10^6^ samples for each posterior density except for PHS and DMA. Because of the singular data, numeric differentiation fails, hence a standard Metropolis-Hastings algorithm was used. Modelling assumptions are described in the respective results sections.

All data is available upon request from the corresponding author

## Author Contributions

TH designed and supervised the project. RD enlisted all control participants, conducted all QST measurements as well as acquired and analyzed all data. VL recruited all control participants. MC provided and recruited all lipedema patients. DT conducted the Bayesian inference. TH, RD, and DT wrote the manuscript. RD, DT, VL, MC, and TH reviewed and edited the manuscript.

## Data Availability

All data produced in the present study are available upon reasonable request to the authors.

## Acknowledgments

We thank all patients and participants for support of the study and providing their respective data. The project was funded by the DFG (459479161). The authors have no conflicts of interest to declare.

